# Ablation STrategies for Repeat PrOcedures in Atrial Fibrillation Recurrences despite Durable Pulmonary Vein Isolation ASTRO - AF Trial

**DOI:** 10.1101/2024.04.09.24305585

**Authors:** Boris Schmidt, Stefano Bordignon, Andreas Metzner, Philipp Sommer, Daniel Steven, Tilmann Dahme, Matthias Busch, Roland Richard Tilz, David Schaack, Andreas Rillig, Christian Sohns, Arian Sultan, Karolina Weinmann-Emhardt, Astrid Hummel, Julia Vogler, Thomas Fink, Jakob Lueker, Alexander Pott, Christian Heeger, KR Julian Chun

**Affiliations:** Cardioangiologisches Centrum Bethanien, Frankfurt, Germany; Universitätsklinikum Frankfurt, Medizinische Klinik 3-Klinik für Kardiologie, Frankfurt, Germany; University Heart and Vascular Center Hamburg (UHZ), Hamburg, Germany; Clinic for Electrophysiology, Herz-und Diabeteszentrum Nordrhein-Westfalen, Ruhr-Universität Bochum, Bad Oeynhausen, Germany; University Hospital Cologne - Heart Center, Cologne, Germany; Uniklinik Ulm, Klinik für Innere Medizin II, Ulm, Germany; Klinikum Esslingen, Klinik für Kardiologie, Angiologie und Pneumologie, Esslingen, Germany; Universitaetsmedizin Greifswald, Greifswald, Germany; Helios Hanseklinikum Stralsund, Klinik für Innere Medizin und Kardiologie, Stralsund, Germany; Schleswig-Holstein University Clinic, Lubeck Campus, Luebeck, Germany; Bonifatius Hospital Lingen, Klinik für Kardiologie und Rhythmologie

**Keywords:** atrial fibrillation, LAA isolation, cryoballoon

## Abstract

**Background:** Ablation strategies for patients with symptomatic atrial fibrillation (AF) and isolated pulmonary veins (PV) vary and their impact on arrhythmia recurrence remains unclear. This prospective randomized German multi—center trial sought to compare two ablation strategies in this patient cohort.

**Methods:** Patients with AF despite durable PV isolation were randomly assigned at seven centers to undergo low-voltage area (LVA) ablation using 3D mapping and irrigated radiofrequency current ablation (group A) or empirical left atrial appendage isolation (LAAI) utilizing the cryoballoon (CB) followed by staged interventional LAA closure (group B). The primary endpoint was freedom from atrial tachyarrhythmias between 91 and 365 days after index ablation. The study was powered for superiority of LAAI compared to LVA.

**Results:** Patients (40% female, mean age 68.8±8 years) with paroxysmal (32%) or persistent AF (68%) were randomized to undergo LVA ablation (n=79) or CB guided LAAI (n=82). After a planned interim analysis enrollment was halted on January 10^th^ 2023.

In the LAAI group 77/82 LAAs were successfully isolated with subsequent LAAC in 57 patients. Procedure related complications occurred in 4 (5%) and 11 (13.5%) patients in group A and B, respectively (P=0.10). The median follow-up was 367 (IQR 359-378) days. The Kaplan Meier point estimate for the freedom from a primary endpoint event was 51.7% (CI 40.9-65.4%) for group A and 55.5% (CI 44.4-69.2%; p=0.8069).

**Conclusions:** The present study did not detect superiority of CB guided LAAI over LVA ablation in patients with AF despite durable PVI.

It was registered at https://clinicaltrials.gov/study/NCT04056390

**Clinical Perspectives:** *What is new?:*

- This is the first randomized multi-center study to compare two different ablation strategies in AF patients with durable PVI.
- Empirical LAAI was not associated with better outcome in comparison to low-voltage area ablation.

*What are the clinical implications?:*

- LAAI should not be advocated as a stand-alone ablation strategy for patients with AF recurrences after prior ablation.
- The patient with AF recurrence after prior catheter ablation should be informed that if all PVs are found durably isolated the optimal ablation strategy remains uncertain.

## Introduction

Despite several improvements, atrial fibrillation (AF) recurrences after a first ablation procedure remain common.(1–3) A considerable number of patients subsequently undergoes repeat procedures for symptomatic AF recurrences, but the optimal ablation strategy is not well investigated and remains ill defined.(4, 5)

In general, uncertainty exists, whether an empirical or an individualized strategy should be employed.(6, 7) In the presence of atrial myopathy, low voltage areas (LVA) unveiled by electroanatomical mapping may be a potential ablation target for an individualized approach.(8, 9) Yet, conflicting data on the value of LVA ablation adjunctive to PVI for first AF ablation procedures was published.(10–12)

Alternatively, empirical ablation such as electrical isolation of the left atrial appendage (LAA) adjunctive to PVI was found to be associated with favorable outcomes after a first AF ablation procedure in selected patients.(13, 14)

Of note, studies investigating the value of ablation in addition to PVI as a first-line approach were confounded by the fact, that non-durable PVI contributes to AF recurrences to some extent.

Therefore, the goal of the study was to compare two contemporary ablation strategies for patients with AF recurrences despite durably isolated PVs in a prospective randomized multi-center study.

## Methods

### Trial Design and Oversight

The trial was approved by the ethics committee of the Landesaerztekammer Hessen (2023-3251-evBO) and complies with the declaration of Helsinki. It was registered at clinical trials.gov (NCT04056390). The study devices were all CE marked. Patients had to sign the patient informed consent form prior to enrollment. The trial was supported by an unrestricted educational grant from Medtronic. The company had no role in the design or execution of the trial or in the preparation of the manuscript. The German health insurance system covered all costs related to catheter ablation. An independent data and safety monitoring board oversaw the trial and reviewed accumulated data.

#### Patients

Patients with symptomatic non-valvular AF despite at least one prior AF ablation attempt were eligible for the trial. The protocol did neither specify for the number nor the type of prior AF ablations. To be eligible for enrollment, patients had to be 18-85 years old. Mild to moderate left atrial enlargement up to a diameter of 55mm was allowed as well as a mildly reduced left ventricular ejection fraction (>45%).

Patients were excluded if they had any contra-indication to repeat catheter ablation or a potentially reversible cause for AF (e.g. hyperthyroidism, severe mitral regurgitation). Patients with chronic obstructive pulmonary disease treated with bronchodilators and patients with obstructive sleep apnea syndrome were excluded.

After informed consent, invasive PV re-mapping was performed. In case of resumed left atrial-to-PV conduction patients were also excluded from the study before randomization (screen failure). A full list of inclusion and exclusion criteria is provided in the supplement table 1.

#### Study course

After obtaining written informed consent, patients were prepared for the ablation procedure according to the clinical standard mandating the assessment of left atrial size, left ventricular ejection fraction, the exclusion of severe valvular dysfunction as well as the exclusion of intracardiac thrombus. After having confirmed durable PVI using a standard circular mapping catheter study group assignment was performed using randomly generated numbers (Study design is depicted in Figure S1 in the Supplement).

#### Catheter Ablation

Ablation procedures were conducted under during deep sedation using midazolam, fentanyl and a continuous infusion of propofol. Vital parameters were continuously monitored. Unfractionated heparin was repeatedly administered to maintain an activated clotting time between 250 and 400s. After single transseptal puncture, a circular mapping catheter was placed in each PV in a sequential fashion to assess PVI. If AF was present at the beginning of the procedure, electrical cardioversion was performed to restore normal sinus rhythm. In case of electrical isolation of all PVs, the patient underwent randomization.

#### Substrate modification (Group A)

Upon confirmation of durable PVI a second transseptal puncture was performed to insert an ablation catheter. A detailed electroanatomical LA map during sinus rhythm was acquired aiming at >500 mapping points. LVA was defined as local voltage of 0.2-0.5 mV, while <0.2mV was considered scar tissue and >0.5mV was considered healthy myocardium. The presence and distribution of LVA was categorized into 6 different left atrial regions: septal, anterior, lateral, inferior, roof and posterior. At LVA irrigated radiofrequency current ablation was performed using a power of 30-40W and a flush rate of 8-30ml/min. The ablation endpoint was local amplitude attenuation of ≥90% at each LVA site. Operators were encouraged to encircle by ablating at the LVA perimeter and ensure loss of capture at 10 milliamp stimulation, and/or completely cover all LVA with ablation lesions. The tagged ablation lesions should confirm encircling and/or covering of the entire LVA indicated by the mapping system. Operators were encouraged to connect 2 neighboring LVAs or anchor LVA to anatomic structure such as the isolated PV or valve annuli to avoid creating slow conduction zones or unanchored islands of LVA that might be deemed to be potentially arrhythmogenic. Bidirectional conduction block was required.

#### Group B: LAA-Isolation

The transseptal sheath was exchanged for a steerable 12F sheath via a guidewire positioned in the left superior PV. Thereafter, a cryoballoon (CB; Arctic Front™, Medtronic, USA) was advanced to the LA and navigated to the LAA using an inner-lumen circular mapping catheter (Achieve™, Medtronic). After confirming complete LAA sealing by the CB using occlusion angiograms cryothermal energy were applied for 300 seconds aiming at LAA isolation. After electrical LAA-isolation a second energy application was performed to consolidate the ablation lesion.

At the end of the procedure all catheters and sheaths were removed and hemostasis was achieved as per center’s standard. Patients underwent transthoracic echocardiography to exclude pericardial effusion. Oral anticoagulation was resumed thereafter.

#### LAA closure

Patients in group B were advised to undergo an interventional LAA closure 4-6 weeks after the index ablation (Figure S1 supplement). This included a LAA re-mapping procedure using a circular mapping catheter. In case of electrical LAA reconnection, repeat LAA isolation using a CB was recommended. In case of durable LAA isolation, LAA closure using a contemporary occluder device was performed under transesophageal echocardiography guidance.

Patients who underwent a second LAA isolation procedure were asked to return for a LAA closure procedure another 4-6 weeks later.

#### Follow-up

Before hospital discharge patients underwent echocardiography to rule out pericardial effusion. At minimum, a 12 lead ECG was obtained. It was recommended to stop all membrane active antiarrhythmic drugs (AAD) before discharge and it was mandatory to stop AADs 90 days (end of the blanking period) after the procedure.

Follow-up included a telephone interview 4 weeks after the procedure to assess adverse events.

At 3, 6 and 12 months post-ablation, outpatient visits including a 72 hour Holter ECG were performed to assess the patient’s arrhythmia status.

In patients with a LAA closure device a transesophageal echocardiogram was performed 6 weeks after implantation to assess LAA closure patency and to rule out device related thrombus.

#### Study endpoints

The primary study endpoint was freedom from documented recurrence of any AF or atrial tachycardia (AT) lasting > 30 seconds between day 91 and 365 after the index procedure.

The safety endpoint was defined as the incidence of peri-procedural complications such as major bleeding requiring intervention, phrenic nerve palsy, pericardial tamponade, thrombembolic events, atrio-to-esophageal fistula or death.

All data was collected, digitally stored and monitored by an independent research organization (CRO Kottmann/RQM+, Hamm, Germany).

#### Statistical analysis

The hypothesis was that LAA isolation was superior to substrate modification for the treatment of AF recurrences despite durably isolated PVs. To test this assumption, a two-sided log rank test for freedom from AF/AT recurrence with a significance level alpha=5% was used. Given historical 12 months freedom from AF/AT rates of 75% after electrical LAA and 55% after substrate modification. This corresponds to a hazard ratio of 0.48. Assuming a slightly larger hazard ratio of 0.55 than observed in this small study, a power of 80% and a significance level of alpha=5%, it was calculated that a total sample size of 256 randomized patients was required. This calculation already assumed a drop-out rate of 10%.

An interim-analysis for safety was planned after randomization of 50% of the study population. Preliminary data on adverse events and freedom from AF/AT as well as overall survival were analyzed and presented to a Data Safety Monitoring Board.

In addition to the results of the interim analysis, the final data set after completion of the follow-up for all patients will be reported below. As of 22st January 2024, the primary end-point status of all randomly assigned patients was known, and follow-up censoring was applied.

Analyses were conducted by an independent biostatistician with the use of the statistical software package SAS@, Version 9.4 under Windows@ XP Statistics software.

## Results

### Patients

Between July 2019 and January 2023, at seven German centers a total of 327 patients were consented to participate in the study. After invasive re-mapping 147 patients had to be excluded for non-durable PVI. Finally, 161/327 screened patients fulfilled all inclusion and exclusion criteria and were subsequently randomized (Figure 1).

**Figure 1:**
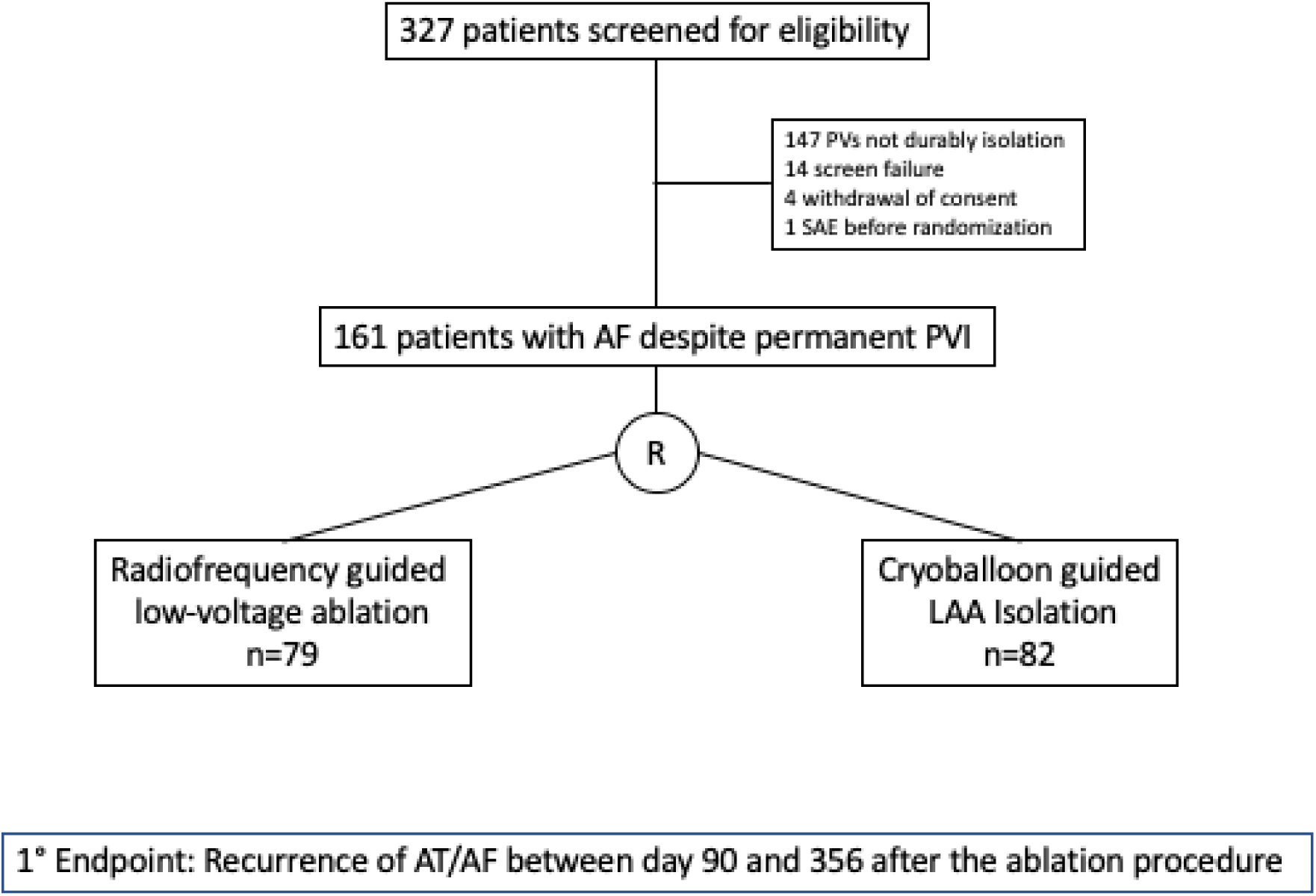
Study flow. AF: Atrial Fibrillation. AT: Atrial Tachycardia. PVI: Pulmonary Vein Isolation. LAA: Left Atrial Appendage.

Details of the patient demographics are given in table 1. Mean age of the study cohort was 68± 8years and 65 patients (40.4%) were female. Patients presented with paroxysmal, persistent and long-standing persistent AF in 31%, 66% and 1 % of cases after a median of 2 (IQR 1-2) prior AF ablation procedures. At the time of enrollment 128 (79.5%) patients were on antiarrhythmic drug therapy. The median time from the last ablation was 19 months (q1-q3: 10-50). The LA diameter was mildly enlarged and mean left ventricular ejection fraction was normal.

**Table 1.** Demographics. BMI: Body Mass Index. AF: Atrial Fibrillation. PAF: Paroxysmal Atrial Fibrillation. TIA: Transient Ischemic Attack. SE: Systemic Embolism. CIED: Chroni implantatble electronic device. RF: Radiofrequency Ablation. CB: Cryoballoon ablation.

|  | Total Cohort (n=161) | Substrate Modification (n=79) | CB guided LAA Isolation (n=82) |
| --- | --- | --- | --- |
| Female Gender (%) | 65 (40%) | 36 (46%) | 29 (35%) |
| Mean Age | 68.6 ± 8 | 67.6 ± 8 | 67.9 ± 8 |
| Median BMI (IQR) | 28 (25-31) | 29 (26-32) | 28 (25-31) |
| Type of AF |  |  |  |
| PAF | 52 (32%) | 32 (41%) | 20 (24%) |
| Persistent AF | 107 (66%) | 46 (58%) | 61 (74%) |
| LS persistent AF | 2 (1%) | 1 (1.3%) | 1 (1.2%) |
| Median time from AF diagnosis | 60 (26-98) | 60 (24-84) | 62 (27-108) |
| Median Time from last AF ablation | 19 (10-50) | 20 (8-48) | 19 (11-52) |
| Arterial Hypertension | 122 (76%) | 63 (80%) | 59 (72%) |
| Diabetes mellitus | 22 (14%) | 13 (16%) | 9 (11%) |
| History of Stroke/TIA/SE | 14 (9%) | 8 (10%) | 6 (7%) |
| Coronary Artery Disease | 32 (20%) | 16 (20%) | 16 (20%) |
| Implanted CIED | 9 (6%) | 5 (6%) | 4 (5%) |
| No. of prior AF ablations |  |  |  |
| 1 | 76 (47%) | 36 (46%) | 40 (49%) |
| 2 | 65 (40%) | 31 (39%) | 34 (41%) |
| 3 | 15 (9%) | 9 (11%) | 6 (7%) |
| 4 | 5 (3%) | 3 (4%) | 2 (2%) |
| Prior Ablation Modality |  |  |  |
| RF | 111 (69%) | 54 (68%) | 57 (70%) |
| CB | 54 (34%) | 24 (30%) | 30 (37%) |
| Other | 18 (11%) | 12 (15%) | 6 (7%) |
| Left Atrial Diameter (mm) | 43 ± 5 | 43 ± 6 | 42 ± 5 |
| Left vemtricular ejection fraction (%) | 58 ± 7 | 58 ± 7 | 58 ± 7 |
| Failed antiarrhythmic drugs | 138 (86%) | 65 (82%) | 73 (89%) |

Patient enrollment was prematurely halted for futility on January 10^th^ 2023 according to the recommendation of the Data Safety and Monitoring Board (Supplement Figure S2).

### Procedural data

In group A substrate modification was performed in 79 patients. Electroanatomical mapping detected at least one low-voltage area in 69/79 (87%) patients. The median number of LVA regions was 2 (range 0-6). The most prevalent LVA locations were in the anterior (61%), posterior (47%), roof (47%) and septal (32%) left atrium. A detailed description of low-voltage area localization is given in figure 2.

**Figure 2.**
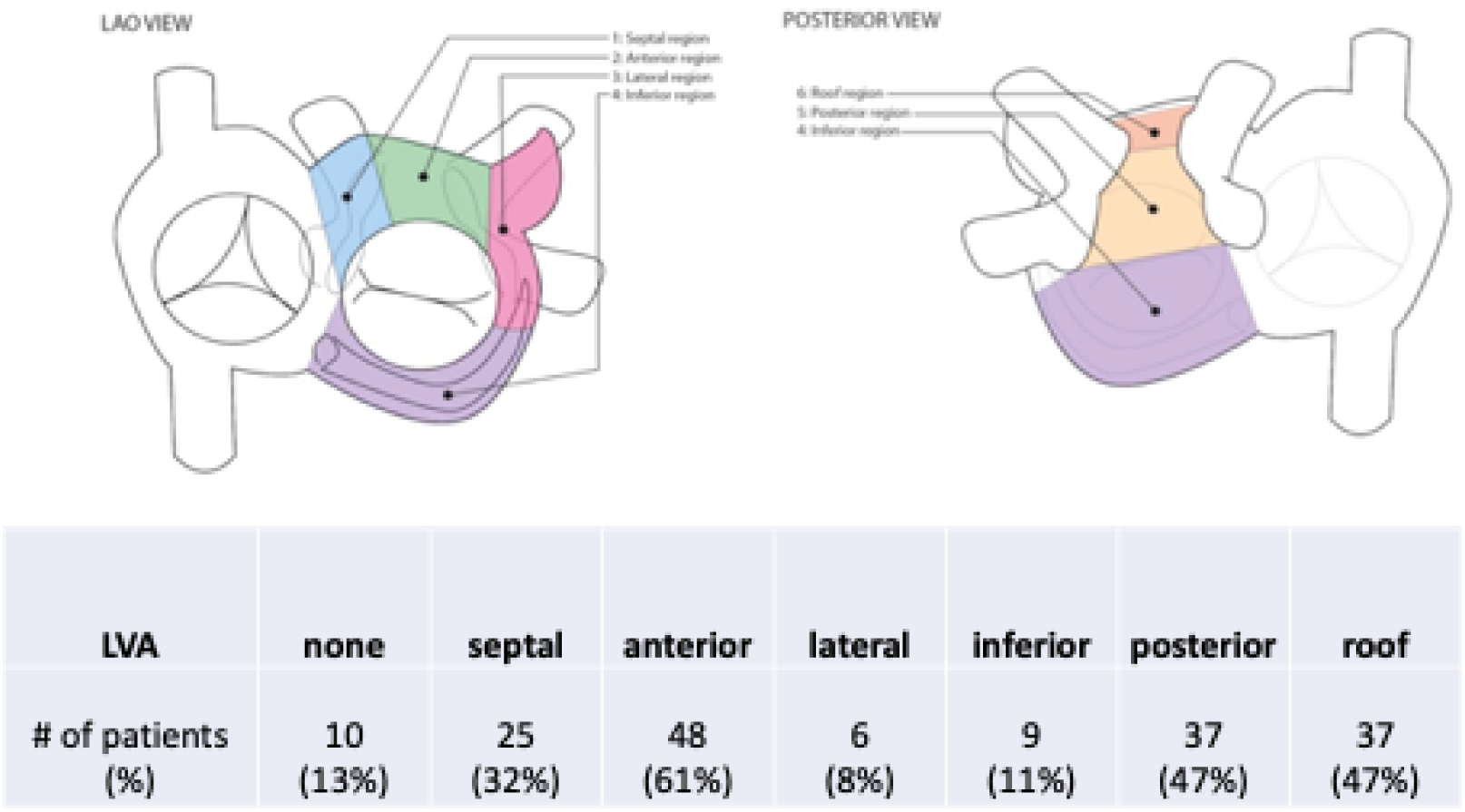
Group A. Representation of low-voltage areas (LVA).

During LVA ablation additional linear lesion ablation was performed in 75 (95%) patients. In 2 (2.5%) patients, the LAA was inadvertently electrically isolated. In 10 patients without LVA linear ablation was carried out in 9 and wide area PV re-ablation in 1, respectively (for details see figure S 3in the supplement).

The median ablation time was 14 (10–26) mins. The mean skin-to-skin procedure and fluoroscopy times were 90±42 minutes and 8.0±4.3 minutes, respectively (Table 2).

**Table 2:** Procedural details. CB: Cryoballoon Ablation. LAA: Left atrial appendage. LVA: low voltage ablation. LAAI: Left atrial appendage isolation. LAAC: left atrial appendage closure

|  | Substrate Modification (n=79) | CB guided LAA Isolation (n=82) |
| --- | --- | --- |
| Number of Procedures | 79 | 163 |
| Median (range) # of Procedures per patient | 1 (1-1) | 2 (1-3) |
| Mean procedure time of index ablation | 90±42 | 70±34 |
| Mean fluoroscopy time of index ablation | 8.0±4.3 | 9.3±7.0 |
| Patients with LVA (%) | 69 (87%) | N/A |
| Patients with linear Ablation (%) | 74 (94%) | 0 |
| LAAI achieved during first ablation | 0 | 77/81 (95.1%) |
| LAAI and LAAC | 0 | 56 (68%) |

In group B, LAA isolation was successfully performed in 77/81 patients (95.1%; Figure 3). One patient was treated by radiofrequency ablation according to group A due to a technical error during the randomization process.

**Figure 3.**
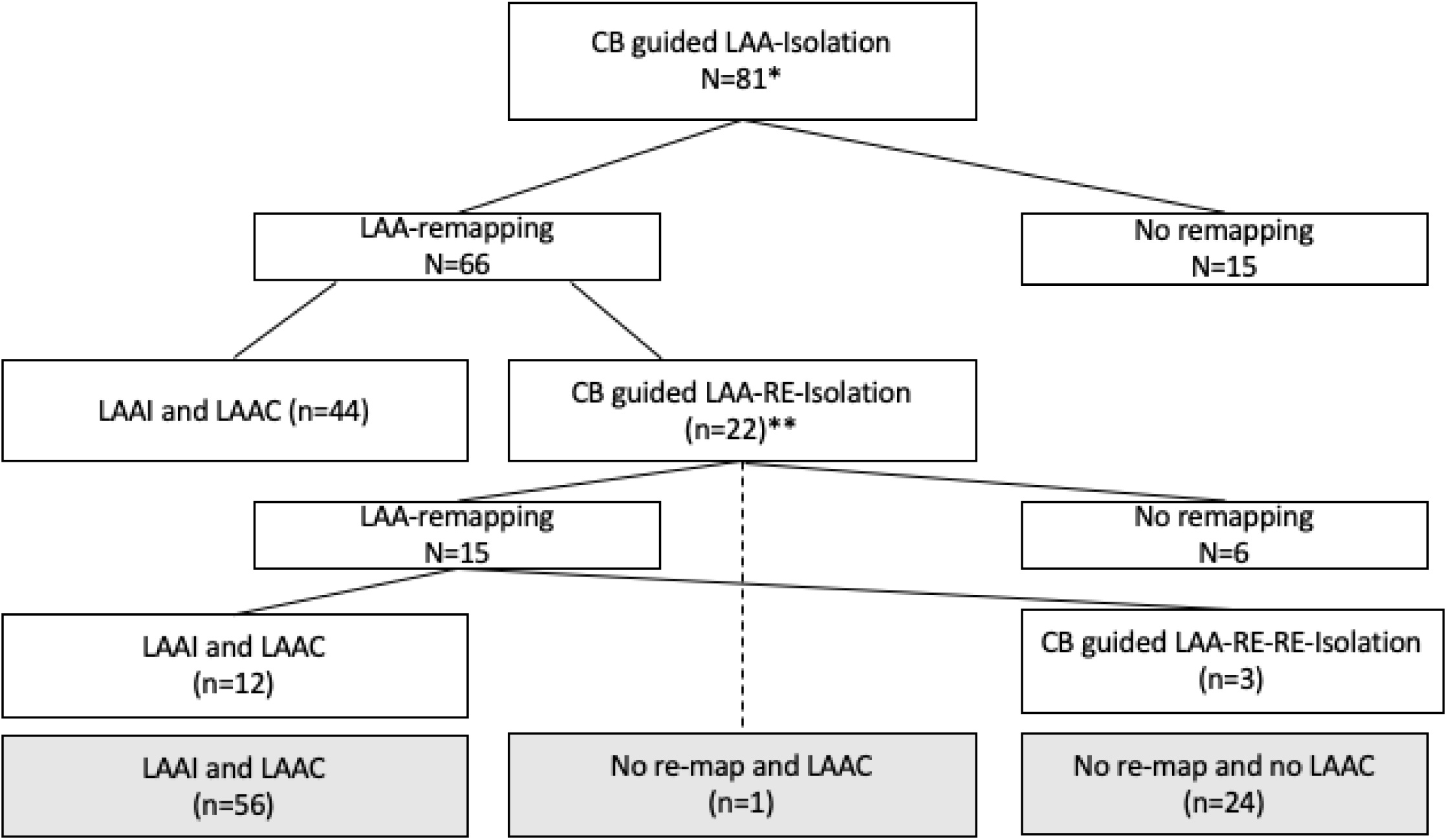
Study flow for patients in group B. * 1 patient did not receive LAA-Iso. ** 1 patient LAAC during LAA-reisolation. CB: cryoballoon, LAA: left atrial appendage. LAAC: left atrial appendage closure.

LAA isolation was achieved with the first CB application in 55 patients (67.9%). In the remaining patients 2, 3,4 and 5 applications were required to achieve isolation in 14, 4, 3 and 1 patients, respectively. The median number of CB applications per patient was 2 (IQR 2-3). The mean skin-to-skin procedure and fluoroscopy times were 70±34 minutes and 9.3±7.0 minutes, respectively.

### Remapping and LAA closure

After a median of 48 (42–59) days 66/81 patients (81.5%) returned for the re-mapping procedure (figure 2). Durable LAA isolation was documented in 44 (67%) patients and interventional LAA closure was performed accordingly.

The remaining 22 patients underwent a repeat LAA isolation procedure. In one of these patients LAA closure was carried out concomitantly during the management of pericardial tamponade. Of these, 14 patients returned for a second re-mapping. In 12 (86%) patients LAA isolation was documented and LAA closure was performed.

In summary, 56/81 patients (69%) had documented LAA isolation followed by closure, in 24 patients LAA isolation status remained unknown and no closure was performed and in a single patient LAA closure was performed concomitantly with a repeat isolation procedure.

### Procedural Complications

In total 15 patients (9.3%) experienced periprocedural serious adverse events with 4 patients (5.0%) from group A and 11 (13.4%) patients from group B (p=0.10; Table 3). Of note, in group A and B 79 and 163 invasive procedures per patient were carried out, respectively.

**Table 3.** Procedural Complications.

|  | Substrate<br>Modification<br>(n=79) | CB guided LAA<br>Isolation and<br>LAAC (n=82) | p-value |
| --- | --- | --- | --- |
| Death | 0 | 0 |  |
| Stroke | 0 | 1 (1.2%) | 1.0 |
| LAA thrombus | 0 | 2 (2.4%) | 0.4970 |
| Pericardial effusion | 2 (2.6%) | 4 (4.9%) | 0.6819 |
| Pericardial effusion<br>requiring<br>intervention | 1 (1.3%) | 4 (4.9%) | 0.3676 |
| Access site<br>complication | 1 (1.3%) | 2 (2.4%) | 1.0 |
| Infection | 1 (1.3%) | 1 (1.2%) | 1.0 |
| Other | 0 | 1 (1.2%) | 1.0 |
| **Total** | **4 (5.0%)** | **11 (13.4%)** | **0.1022** |

No ablation related death was reported. Pericardial effusion occurred in 2 (2.5%) and 4 (4.9%) patients in group A and B, respectively (p=0.68). While only 1 pericardial effusion from group A required subxiphoidal drainage, this applied to all 4 in group B. In the latter group, all pericardial effusions were attributable to the ablation procedure.

Vascular access complications requiring medical intervention were observed in one patient and 2 patients from group A and B, respectively (p=1.0). After LAA isolation, in 2 patients a LAA thrombus was detected. In one patient, the thrombus resolved with vitamin K antagonist therapy. Subsequently LAA closure was performed. In the second patient LAA thrombus persisted despite oral anticoagulation but the patient refused interventional treatment.

### Primary Endpoint

The median follow-up was 367 (IQR 359-378) days. After completion of the follow-up for all randomized patients, a final analysis was performed. During the course of the study, 11 and 3 patients withdrew consent or were lost to follow-up (6 and 8 from group A and B, respectively). Thus, the efficacy analysis was computed for 147 patients. It showed, that 35 and 33 patients from group A and group B reached the primary endpoint. The Kaplan Meier point estimate for the freedom from a primary endpoint event was 51.7% (CI 40.9-65.4%) for group A and 55.5% (CI 44.4-69.2%) (p=0.8069; Figure 4, Central Illustration). More specifically, the point estimate rates of recurrent AF were 38.8% (CI 25.2-49.9%) versus 41.9% (CI 28.3-53.0%; p=0.50) and of recurrent AT 15.8% (CI 6.8-24.0%) and 7.0% (CI 0.9-12.8%; p=0.11) in group A and group B, respectively.

**Figure 4:**
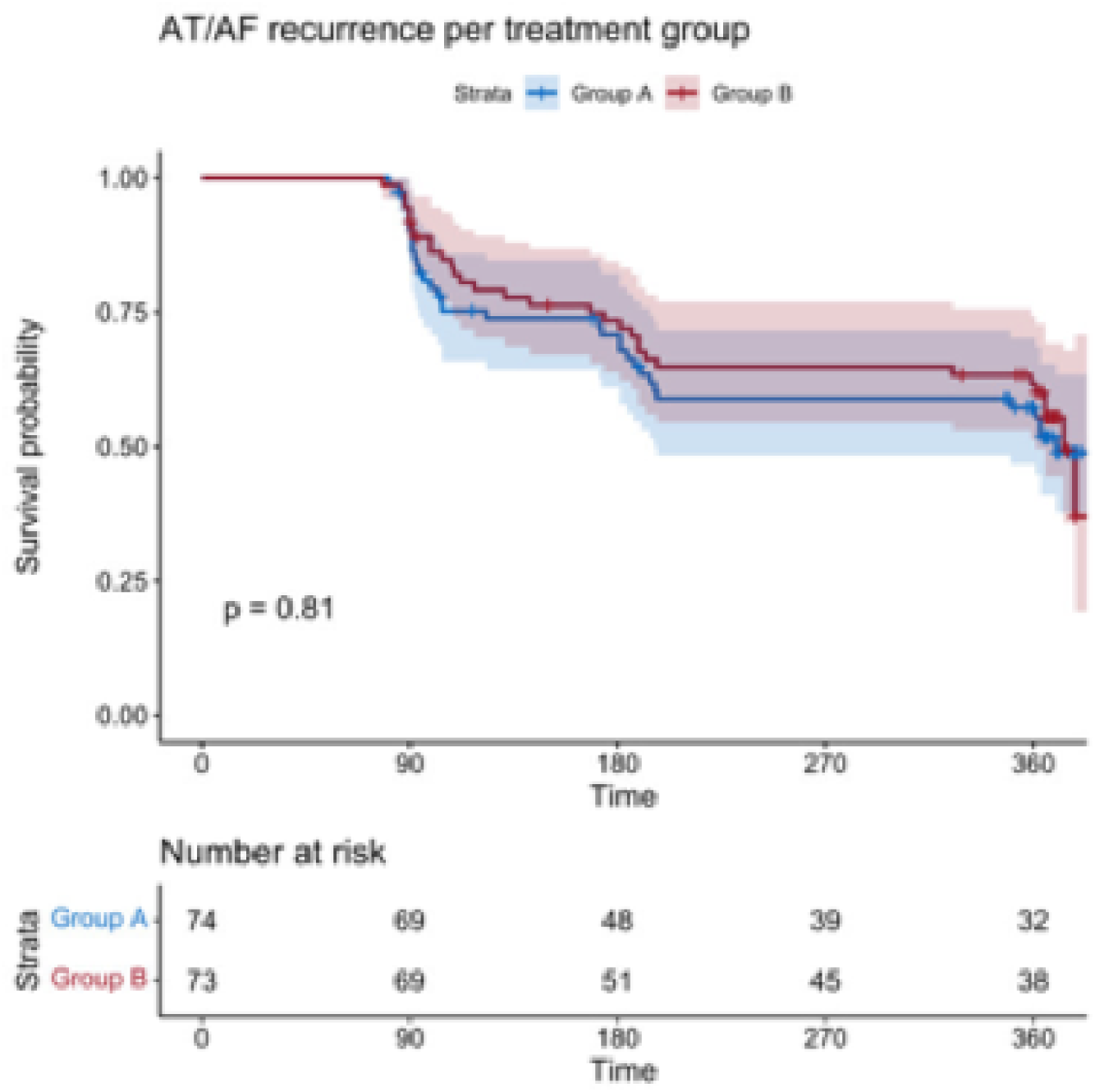
Kaplan Meier Analysis for the Primary Endpoint.

### Serious adverse events during follow-up

One patient from each group died during follow-up 284 and 297 days after the ablation. Both deaths were not related to the study procedure.

In one patient a pericardial effusion requiring drainage occurred 8 months after the LAA closure procedure. Computed tomography imaging did not show any device erosion. In 2/5 patients repeat subxiphoidal drainage was required for recurrent pericardial effusion after an initial drainage (see above).

In one patient from group B a stroke occurred 12 days after the LAA closure procedure. Imaging revealed an embolic stroke to the middle cerebral artery.

Major bleeding occurred in one patient of group B and was related to a newly diagnose colorectal cancer.

### Drug therapy during follow-up

During follow-up 86.3% of patients remained on oral anticoagulation (Figure 5). Oppositely, in group B, 48.5% of patients were still taking oral anticoagulation and 53% had either reduced the antithrombotic regimen to either single antiplatelet or no therapy.

**Figure 5:**
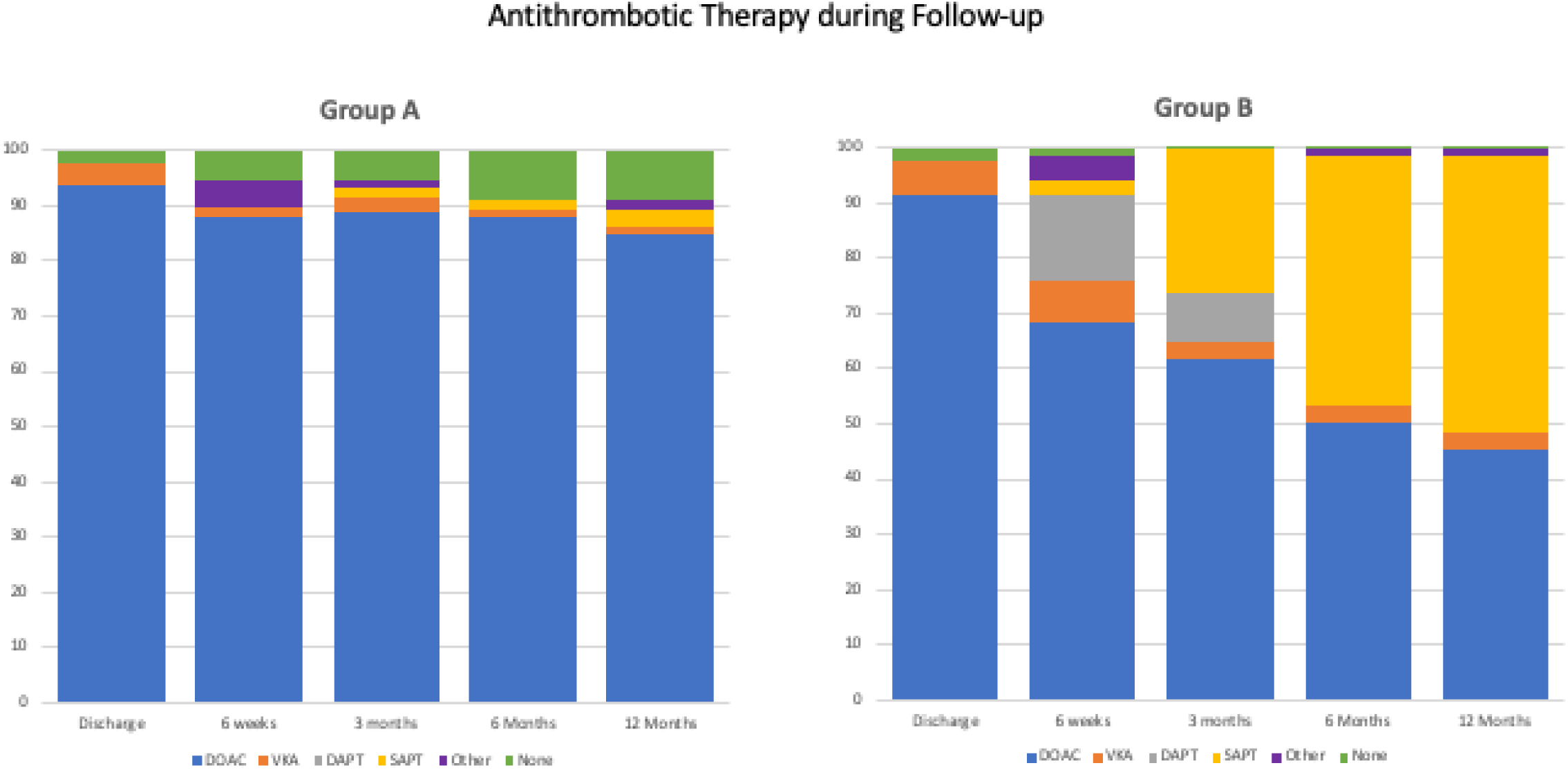
Overview of the antithrombotic therapy during the study for the two groups. DOAC: Direct oral anticoagulant. VKA: Vitamin K antagonist. DAPT: dual antiplatelet therapy, SAPT: single antiplatelet therapy. Central Illustration: Study overview. In total, 161 patients with atrial fibrillation (AF) despite durable pulmonary vein isolation (PVI) were randomized at seven German centers to undergo cryoballoon (CB) guided left atrial appendage isolation (LAAI) or ablation of low-voltage areas (LVA) in the left atrium (LA). Freedom from atrial tachyarrhythmias was similar after 12 months.

At the 12 months follow-up, 9.2 and 24.7% of patients in group A and B were taking class I or III antiarrhythmic drugs.

## Discussion

In the present prospective randomized multi-center trial LAA isolation using the CB did not improve the freedom from AT/AF recurrence in patients with AF despite durably isolated pulmonary veins in comparison to LVA ablation. To the best of our knowledge this is the first randomized trial carried out in patients with proven PVI.

Electrical PVI remains the only unequivocally accepted endpoint during AF catheter ablation.(15) Most studies assessing the optimal strategy during repeat AF ablation include patients with re-connected PVs. Thus far, none of the additional empirical ablation sets including posterior wall isolation and left atrial linear ablation yielded positive results.(16, 17)

Given the improved durability results with modern technology, ablation strategies beyond PVI become increasingly important. In this study, an individualized concept targeting LVA as a potential substrate for AF initiation and perpetuation was compared to an empirical anatomical approach.

### Left Atrial Appendage Isolation

The concept of an empirical ablation applicable to all patients with an unequivocal procedural endpoint seems desirable in terms of generalizability and reproducibility.

The first randomized study investigating the effect of LAAI in addition to PVI and LA substrate ablation was carried out in patients with long-standing persistent AF.(13) In the LAAI group a success rate defined as arrhythmia free survival 24 months after the index ablation was reported to be 76%.

In the present study, LAAI did not lead to better arrhythmia free outcome than the individualized LVA ablation concept. While the acute procedural endpoint was achieved in 95% of cases, CB guided LAAI was associated with a considerable number of recovered LA to LAA conduction (33% of all re-mapped patients) necessitating repeat ablation procedures. Moreover, permanent LAAI eliminates the mechanical LAA function leading to reduced flow velocities, hence increasing the risk for thrombus formation and stroke despite continued OAC.(18, 19) Based on the findings of prior studies, LAAC was recommended to all patients in the LAAI group in case of documented LAAI.(20) Successful LAAC was performed in 69% of the patient population.

Per protocol, patients in group B had to undergo significantly more procedures than patients in the LVA ablation arm partly explaining the trend towards a higher incidence of peri-procedural complications. In particular, the rate of ablation related pericardial effusion requiring intervention was considerable at 4.9%.

On the other hand, patients after LAA closure were less likely to use oral anticoagulation. This may translate into less bleeding complications during long-term follow-up. Prospective randomized strategies comparing oral anticoagulation with LAA closure after ablation are currently underway.(21)

Altogether, it seems prudent to reserve LAAI to patients with proven AF triggers.

### Low-Voltage Area Ablation

LVA ablation is an individualized ablation concept based on the high-density mapping results. Earlier studies have demonstrated, that the presence and extent of atrial fibrosis and LVA correlate not only with ablation outcomes but also with adverse clinical events such as stroke, heart failure and death.(22, 23) In contrast, prospectively randomized studies investigating the value of LVA ablation in addition to PVI reported conflicting data. In the most recent ERASE AF study additional LVA ablation improved AF/AT survival by 38%. In contrast, in a similar patient population, investigators of the STABLE-SR-II study did not find better rhythm outcomes after LVA ablation.

The results of both studies may be confounded by the quality of PV isolation, i.e. the true contribution of LVA versus PV triggers may not be assessed with that study design. In the present study, however, only patients with AF despite already permanently isolated PVs were randomized and the observed effects are solely attributable to LVA ablation. Of note, in the vast majority of cases additional LA linear lesions were deployed to connect LVA areas in an attempt to prevent LA macro-reentrant ATs. In contrast to the application of empirical linear ablation as performed in the STAR AF 2 study, the present linear ablation was carried out in regions of low-voltage increasing the likelihood of bidirectional conduction block.(24) This may have contributed to a relatively low incidence of AT after LVA in group A.

### Study Limitations

The study design compared only two existing ablation strategies with one empirical trigger eliminating approach and one individualized approach. It may be speculated that other strategies such as posterior wall isolation, complete trigger elimination etc. would have improved outcomes, however, in the latest EHRA/APHRS/HRS/LAHRS Expert Consensus Statement on Catheter and Surgical Ablation of AF it was stated that the value of these strategies remain an area of uncertainty.

In the present study, the CB was used off-label to achieve LAAI. This was associated with a considerable cardiac perforation rate and a less than expected efficacy, partially explained by reconnections. In earlier observational studies, complication rate was lower and the outcome was remarkable at 86% of patients free from atrial arrhythmias.(14) One may speculate that, improved devices (shorter, atraumatic tip, re-designed sheath for better co-axiality) may enhance outcome.

Several observational studies have investigated the feasibility of concomitant AF ablation and LAAC in a single procedure.(25, 26) Whilst, this would potentially reduce the procedural risk (less procedures), the feasibility of concomitant LAAI and LAAC has not been assessed systematically. The issue of electrical LAA to LA re-connection and subsequent need for re-ablation may be more difficult to address after LAAC

In this study, no detailed analysis of the right atrial substrate was performed which may of course contribute to AF genesis. We tried to minimize this confounder by excluding patients with lung disease.

The follow-up strategy did not use continuous monitoring but 72-hour Holter recordings at 3 different time points. Thus, the true incidence of the primary endpoint may have been underestimated and AF burden was not assessible.

## Conclusion

The present study did not demonstrate a superiority of CB guided LAA isolation over radiofrequency guided LVA in patients with AF recurrences despite durable PVI. The study was prematurely halted for futility after randomization of 63% of the planned patient population.

## Data Availability

Data can be accessed with permission of the author

## Acknowledgements

We thank the members of the Data Safety and Monitoring Board, Dr. Melanie Gunawardene, Hamburg, Germany; Dr. Laura Perrotta, Firenze, Italy and Dr. Shibu Matthew, Essen, Germany for their careful analysis of the data.

## References

1. Mont L, Bisbal F, Hernández-Madrid A, et al. Catheter ablation vs. antiarrhythmic drug treatment of persistent atrial fibrillation: A multicentre, randomized, controlled trial (SARA study). Eur. Heart J. 2014;35:501–507.

2. Kuck K-H, Brugada J, Fürnkranz A, et al. Cryoballoon or Radiofrequency Ablation for Paroxysmal Atrial Fibrillation. N. Engl. J. Med. 2016;374:2235–45.

3. Poole JE, Bahnson TD, Monahan KH, et al. Recurrence of Atrial Fibrillation After Catheter Ablation or Antiarrhythmic Drug Therapy in the CABANA Trial. J. Am. Coll. Cardiol. 2020;75:3105–3118.

4. Zhou L, He L, Wang W, et al. Effect of repeat catheter ablation vs. antiarrhythmic drug therapy among patients with recurrent atrial tachycardia/atrial fibrillation after atrial fibrillation catheter ablation: data from CHINA-AF registry. Europace 2023;25:382–389.

5. Andrade JG, Deyell MW, Macle L, et al. Healthcare utilization and quality of life for atrial fibrillation burden: the CIRCA-DOSE study. Eur. Heart J. 2023;44:765–776.

6. Hindricks G, Potpara T, Dagres N, et al. 2020 ESC Guidelines for the diagnosis and management of atrial fibrillation developed in collaboration with the European Association for Cardio-Thoracic Surgery (EACTS). Eur. Heart J. 2020:1–126.

7. Brahier MS, Friedman DJ, Bahnson TD, Piccini JP. Repeat Catheter Ablation for Atrial Fibrillation. Hear. Rhythm 2024.

8. Kircher S, Arya A, Altmann D, et al. Individually tailored vs. standardized substrate modification during radiofrequency catheter ablation for atrial fibrillation: a randomized study. EP Eur. 2018;20:1766–1775.

9. Rolf S, Kircher S, Arya A, et al. Tailored Atrial Substrate Modification Based on Low-Voltage Areas in Catheter Ablation of Atrial Fibrillation. Circ. Arrhythmia Electrophysiol. 2014;7:825–833.

10. Huo Y, Gaspar T, Schönbauer R, et al. Low-Voltage Myocardium-Guided Ablation Trial of Persistent Atrial Fibrillation. NEJM Evid. 2022;1.

11. Yang G, Zheng L, Jiang C, et al. Circumferential Pulmonary Vein Isolation Plus Low-Voltage Area Modification in Persistent Atrial Fibrillation: The STABLE-SR-II Trial. JACC Clin. Electrophysiol. 2022;8:882–891.

12. Yang B, Jiang C, Lin Y, et al. STABLE-SR (Electrophysiological substrate ablation in the left atrium during sinus rhythm) for the treatment of nonparoxysmal atrial fibrillation: A prospective, multicenter randomized clinical trial. Circ. Arrhythmia Electrophysiol. 2017;10.

13. Di Biase L, Burkhardt JD, Mohanty P, et al. Left Atrial Appendage Isolation in Patients With Longstanding Persistent AF Undergoing Catheter Ablation. J. Am. Coll. Cardiol. 2016;68:1929–1940. A

14. Yorgun H, Canpolat U, Kocyigit D, Çöteli C, Evranos B, Aytemir K. Left atrial appendage isolation in addition to pulmonary vein isolation in persistent atrial fibrillation: One-year clinical outcome after cryoballoon-based ablation. Europace 2017;19:758–768.

15. Calkins H, Hindricks G, Cappato R, et al. 2017 HRS/EHRA/ECAS/APHRS/SOLAECE expert consensus statement on catheter and surgical ablation of atrial fibrillation: Executive summary. Europace 2018;20:157–208.

16. Kim D, Yu HT, Kim TH, et al. Electrical Posterior Box Isolation in Repeat Ablation for Atrial Fibrillation: A Prospective Randomized Clinical Study. JACC. Clin. Electrophysiol. 2022;8:582–592.

17. Fichtner S, Sparn K, Reents T, et al. Recurrence of paroxysmal atrial fibrillation after pulmonary vein isolation: is repeat pulmonary vein isolation enough? A prospective, randomized trial. Europace 2015;17:1371–1375.

18. Rillig A, Tilz RR, Lin T, et al. Unexpectedly High Incidence of Stroke and Left Atrial Appendage Thrombus Formation After Electrical Isolation of the Left Atrial Appendage for the Treatment of Atrial Tachyarrhythmias. Circ. Arrhythmia Electrophysiol. 2016;9:e003461.

19. Heeger CH, Rillig A, Geisler D, et al. Left Atrial Appendage Isolation in Patients Not Responding to Pulmonary Vein Isolation: Benefit and Risks. Circulation 2019;139:712–715.

20. Zender N, Weise FK, Bordignon S, et al. Thromboembolism after electrical isolation of the left atrial appendage: a new indication for interventional closure? Europace 2019;21:1502–1508.

21. Wazni OM, Boersma L, Healey JS, et al. Comparison of anticoagulation with left atrial appendage closure after atrial fibrillation ablation: Rationale and design of the OPTION randomized trial. Am. Heart J. 2022;251:35–42.

22. Marrouche NF, Wilber D, Hindricks G, et al. Association of atrial tissue fibrosis identified by delayed enhancement MRI and atrial fibrillation catheter ablation: the DECAAF study. JAMA 2014;311:498–506.

23. Masuda M, Matsuda Y, Uematsu H, et al. Prognostic impact of atrial cardiomyopathy: Long-term follow-up of patients with and without low-voltage areas following atrial fibrillation ablation. Hear. Rhythm 2023.

24. Verma A, Jiang C, Betts TR, et al. Approaches to catheter ablation for persistent atrial fibrillation. N. Engl. J. Med. 2015;372:1812–22.

25. Fassini G, Conti S, Moltrasio M, et al. Concomitant cryoballoon ablation and percutaneous closure of left atrial appendage in patients with atrial fibrillation. Europace 2016;18:1705–1710.

26. Phillips KP, Romanov A, Artemenko S, et al. Combining left atrial appendage closure and catheter ablation for atrial fibrillation: 2-year outcomes from a multinational registry. Europace 2020;22:225–231.

